# Precision Oncology Through Dialogue: AI-HOPE-RTK-RAS Integrates Clinical and Genomic Insights into RTK-RAS Alterations in Colorectal Cancer

**DOI:** 10.1101/2025.06.06.25329155

**Authors:** Ei-Wen Yang, Brigette Waldrup, Enrique Velazquez-Villarreal

## Abstract

**Introduction:** The RTK-RAS signaling cascade is a central axis in colorectal cancer (CRC) pathogenesis, governing cellular proliferation, survival, and therapeutic resistance. Somatic alterations in key pathway genes—including KRAS, NRAS, BRAF, and EGFR—are pivotal to clinical decision-making in precision oncology. However, the integration of these genomic events with clinical and demographic data remains hindered by fragmented resources and a lack of accessible analytical frameworks. To address this challenge, we developed AI-HOPE-RTK-RAS, a domain-specialized conversational artificial intelligence (AI) system designed to enable natural language– based, integrative analysis of RTK-RAS pathway alterations in CRC.

**Methods:** AI-HOPE-RTK-RAS employs a modular architecture combining large language models (LLMs), a natural language-to-code translation engine, and a backend analytics pipeline operating on harmonized multi-dimensional datasets from cBioPortal. Unlike general-purpose AI platforms, this system is purpose-built for real-time exploration of RTK-RAS biology within CRC cohorts. The platform supports mutation frequency profiling, odds ratio testing, survival modeling, and stratified analyses across clinical, genomic, and demographic parameters. Validation included reproduction of known mutation trends and exploratory evaluation of co-alterations, therapy response, and ancestry-specific mutation patterns.

**Results:** AI-HOPE-RTK-RAS enabled rapid, dialogue-driven interrogation of CRC datasets, confirming established patterns and revealing novel associations with translational relevance. Among early-onset CRC (EOCRC) patients exhibited a significantly lower prevalence of RTK-RAS alterations compared to late-onset disease (67.97% vs. 79.9%; OR = 0.534, p = 0.014), suggesting the involvement of alternative oncogenic drivers. In KRAS-mutant patients receiving Bevacizumab, early-stage disease (Stage I–III) was associated with superior overall survival relative to Stage IV (p = 0.0004). In contrast, BRAF-mutant tumors with microsatellite-stable (MSS) status displayed poorer prognosis despite higher chemotherapy exposure (OR = 7.226, p < 0.001; p = 0.0000). Among EOCRC patients treated with FOLFOX, RTK-RAS alterations were linked to worse outcomes (p = 0.0262). The system also identified ancestry-enriched noncanonical mutations—including CBL, MAPK3, and NF1—with NF1 mutations significantly associated with improved prognosis (p = 1 × 10⁻□).

**Conclusions:** AI-HOPE-RTK-RAS exemplifies a new class of conversational AI platforms tailored to precision oncology, enabling integrative, real-time analysis of clinically and biologically complex questions. Its ability to uncover both canonical and ancestry-specific patterns in RTK-RAS dysregulation—especially in EOCRC and populations with disproportionate health burdens—underscores its utility in advancing equitable, personalized cancer care. This work demonstrates the translational potential of domain-optimized AI tools to accelerate biomarker discovery, support therapeutic stratification, and democratize access to multi-omic analysis.

## Introduction

Colorectal cancer (CRC) is a leading cause of cancer-related morbidity and mortality globally, with an increasing incidence of early-onset CRC (EOCRC)—diagnosed before age 50—particularly among high-risk populations [1–5]. While the molecular landscape of CRC is complex, the receptor tyrosine kinase (RTK)-RAS signaling pathway has emerged as a central driver of tumorigenesis and therapeutic resistance [6–8]. However, characterizing RTK-RAS pathway dysregulation in EOCRC, particularly among different populations, remains limited by data fragmentation in genomic datasets, and a lack of user-friendly analytical tools that integrate clinical and genomic data [5, 9–14]

The RTK-RAS pathway governs essential processes including cell proliferation, survival, and differentiation, and is frequently altered in CRC [5, 17]. Mutations in key genes such as *KRAS*, *NRAS*, and *BRAF* are among the most common genetic alterations in CRC, with *KRAS* mutations occurring in approximately 40% of cases [5, 17]. These alterations play a critical role in clinical decision-making, as they confer resistance to anti-EGFR therapies—one of the primary targeted treatment strategies in metastatic CRC [17–22]. Although *KRAS* and *NRAS* mutations are reportedly more frequent in specific populations [24–25], emerging evidence suggests that the mutation patterns may vary by ancestry. Specifically, recent work by our group and others indicates that RTK-RAS pathway alterations may be less prevalent in EOCRC among ancestry-specific subgroups, with an enrichment of mutations in *CBL*, *NF1*, and *MAPK3* instead [5, 26].

Despite the clinical importance of RTK-RAS pathway alterations, most existing bioinformatics platforms such as cBioPortal [27] and UCSC Xena [28] are built on static user interfaces and require multi-step analysis pipelines, limiting accessibility for non-programmers and hindering rapid translational discovery. These limitations are particularly pronounced when evaluating population-specific differences, pathway co-mutations, or survival outcomes in precision oncology contexts.

Recent breakthroughs in artificial intelligence (AI), particularly large language models (LLMs), now enable natural language–driven bioinformatics pipelines that can translate human queries into executable code [29–34]. While early AI platforms have demonstrated the potential to streamline multi-omic data analysis [35–40], few are purpose-built to interrogate specific pathways like RTK-RAS or to support the integration of genomic and clinical data across CRC cohorts.

To address this gap, we developed AI-HOPE-RTK-RAS (Artificial Intelligence agent for High-Optimization and Precision Medicine focused on RTK-RAS), a conversational AI system designed to investigate RTK-RAS pathway alterations in CRC using integrative, natural language–driven bioinformatics. The platform enables intuitive analysis of mutation frequencies, survival outcomes, treatment resistance patterns, and population-level stratification. In this study, we (1) developed and deployed AI-HOPE-RTK-RAS to analyze public CRC datasets, (2) validated its analytical capabilities by reproducing key trends from RTK-RAS–focused studies, and (3) demonstrated its potential for novel discovery in EOCRC, including ancestry-informed mutation enrichment and prognostic evaluation. Together, these efforts establish AI-HOPE-RTK-RAS as a scalable and accessible tool for RTK-RAS–driven precision oncology research.

## Methods

### Overview of AI-HOPE-RTK-RAS Platform

AI-HOPE-RTK-RAS is a specialized conversational artificial intelligence (AI) system developed to facilitate precision oncology investigations centered on RTK-RAS signaling in CRC. Designed for real-time interaction, the platform allows researchers and clinicians to explore large-scale genomic and clinical datasets through intuitive, plain-language prompts (Figure 1). The system dynamically translates these queries into executable code, performs on-demand bioinformatics analysis, and delivers interpretive outputs relevant to mutation frequency, treatment response, and survival outcomes.

**Figure 1.**
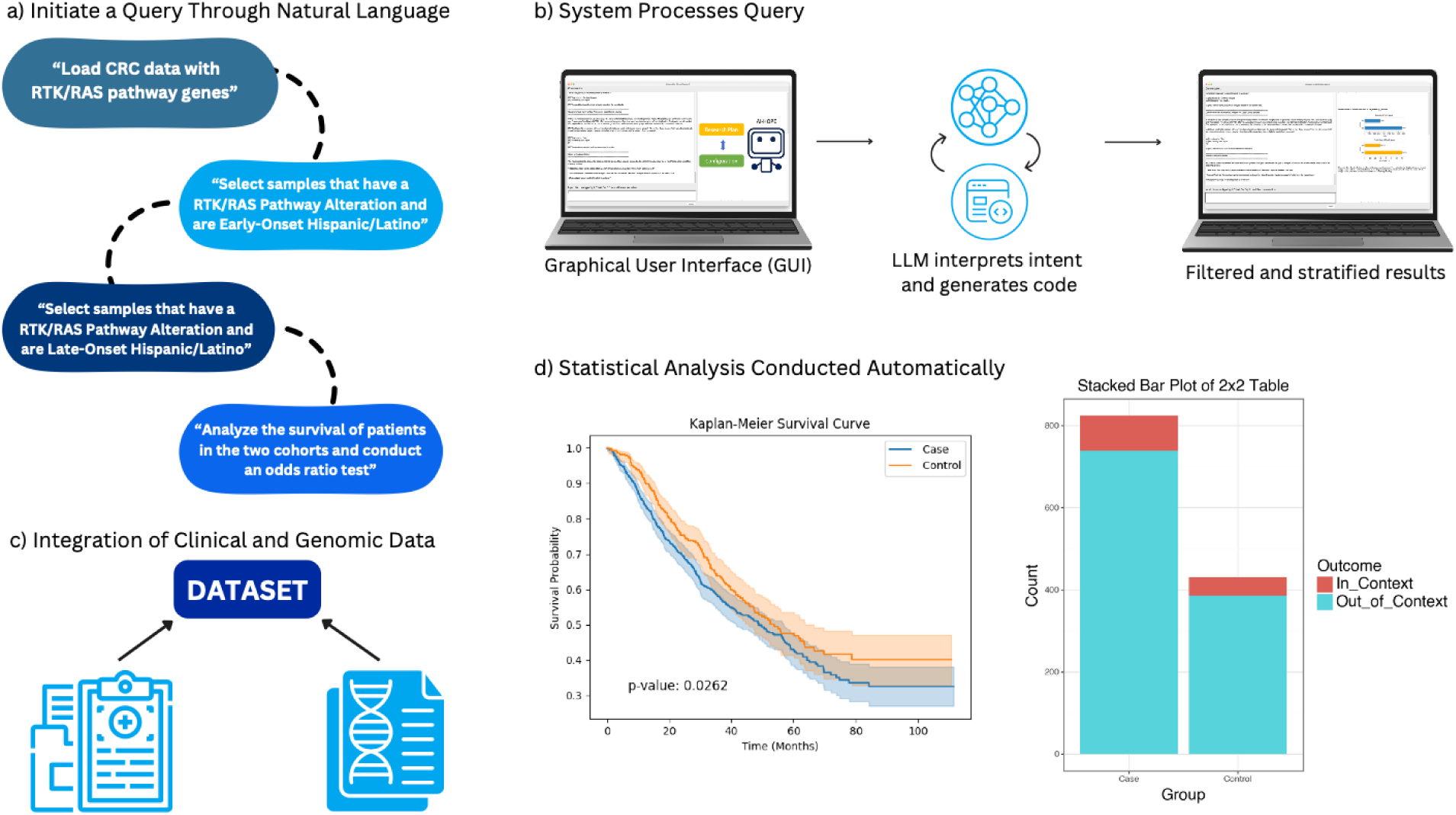
AI-HOPE-RTK-RAS conversational workflow for integrative clinical and genomic analysis. This figure presents the functional pipeline of AI-HOPE-RTK-RAS, an interactive artificial intelligence platform tailored for exploring RTK-RAS signaling in colorectal cancer (CRC). a) The process begins with the user posing a research question in natural language—for example, assessing differences in survival between early- and late-onset Hispanic/Latino patients with RTK-RAS pathway mutations. b) The query is handled through a user-friendly graphical interface, where a large language model (LLM) interprets the request, translates it into executable code, and defines the parameters for subgroup comparison. c) The platform accesses harmonized clinical and genomic datasets—such as those from TCGA and cBioPortal—focusing on RTK-RAS genes including KRAS, NRAS, BRAF, EGFR, ERBB2, CBL, MAPK3, and NF1. Relevant clinical filters (e.g., age, ancestry, treatment exposure) are applied based on the query intent. d) Statistical analyses are carried out automatically, generating results such as survival curves and odds ratios. Outputs are rendered in publication-ready formats alongside narrative interpretations, enabling efficient exploration of complex CRC datasets through a precision oncology lens.

### Data Sources and Curation

Publicly available datasets were obtained from The Cancer Genome Atlas (TCGA) and the cBioPortal for Cancer Genomics, with a focus on colorectal adenocarcinomas. The datasets included somatic mutation profiles, demographic attributes (age, sex, race/ethnicity), clinical variables (tumor location, MSI status, staging), and treatment history, particularly anti-EGFR exposure. Genes of interest within the RTK-RAS pathway included KRAS, NRAS, BRAF, EGFR, ERBB2, CBL, MAPK3, and NF1. All data were reformatted into harmonized, analysis-ready tables using consistent sample identifiers. Controlled vocabularies (e.g., OncoTree, SNOMED) were applied to align diagnostic and phenotypic metadata.

### Natural Language Interface and Query Handling

The user interface is powered by a fine-tuned large language model (LLaMA 3), enabling flexible dialogue-based interaction. Users pose queries in natural language, such as “Show mutation prevalence of KRAS in EOCRC versus LOCRC,” or “Compare survival outcomes for patients with BRAF V600E mutations receiving EGFR inhibitors.” The platform interprets the query intent, checks for ambiguity, and guides users toward analyzable questions when clarification is needed. Complex filtering—such as stratifying by age, ancestry, mutation status, or therapy—is handled seamlessly without requiring programming knowledge.

### Backend Analysis Pipeline

AI-HOPE-RTK-RAS conducts statistical analyses using an integrated Python-based computational core. For binary or categorical variables, Fisher’s exact test and chi-square test are used to assess group differences, and odds ratios are calculated with 95% confidence intervals. Continuous variables are summarized using standard descriptive statistics. Survival analyses utilize Kaplan–Meier estimation with log-rank tests for comparisons; multivariate Cox proportional hazards models are applied for adjusting covariates. The system supports mutation co-occurrence analysis, therapeutic stratification, and ancestry-specific subgroup modeling.

### System Infrastructure and Error Handling

To ensure transparency and reproducibility, the platform employs structured prompting strategies and rule-based logic layers that constrain the output to valid biomedical operations. A retrieval-augmented generation (RAG) component provides access to curated reference materials—such as drug-gene interaction databases and pathway annotations—to enhance the interpretability of results and reduce model hallucinations. Ambiguous queries trigger clarification loops, and all executed commands are logged with versioned output.

### Validation Strategy

To validate platform functionality, we replicated known findings from prior RTK-RAS CRC literature, including mutation frequencies of KRAS, NRAS, and BRAF, and their association with resistance to EGFR-targeted therapies [17–22]. Additionally, we assessed previously reported RTK-RAS mutation profiles among patients with EOCRC from populations with disproportionate health burdens, including the enrichment of noncanonical alterations such as CBL and NF1 [5].

### Comparative Usability Testing

We compared AI-HOPE-RTK-RAS with conventional tools such as cBioPortal and UCSC Xena for usability and analytical throughput. Performance metrics included query response time, flexibility in subgroup definition, and the ability to execute multi-parameter analyses (e.g., filtering by both age and ethnicity). The conversational interface outperformed traditional interfaces in efficiency and reduced the need for specialized bioinformatics skills.

### Output Delivery and Visualization

Final outputs are returned as structured analytical reports, featuring cleanly formatted tables, frequency plots, survival curves, and forest plots. Visualizations are generated using Matplotlib and Plotly libraries with export-quality resolution. Each result is accompanied by a text summary that contextualizes the findings with supporting literature, enabling immediate interpretation and downstream reporting.

## Results

The AI-HOPE-RTK-RAS platform enabled flexible, real-time interrogation of CRC datasets using natural language input to uncover clinically and biologically relevant insights into RTK-RAS pathway dysregulation. Through integrated cohort filtering by mutation status, treatment regimen, tumor stage, microsatellite instability, and demographic features, the system supported automated statistical analysis and visualization, producing results that both confirmed known patterns and revealed new associations with potential translational relevance—particularly within EOCRC and populations with disproportionate health burdens.

### RTK-RAS Alterations in EOCRC by Ancestry

A demographic-stratified analysis compared RTK-RAS pathway mutation frequencies between early- and late-onset CRC patients. Among those under 50 years old, RTK-RAS alterations were detected in 67.97% of cases, versus 79.9% in older counterparts. This difference yielded an odds ratio of 0.534 (p = 0.014), suggesting a reduced prevalence of canonical RTK-RAS alterations in EOCRC, potentially implicating alternative drivers in tumorigenesis in this subgroup (Figure 2).

**Figure 2.**
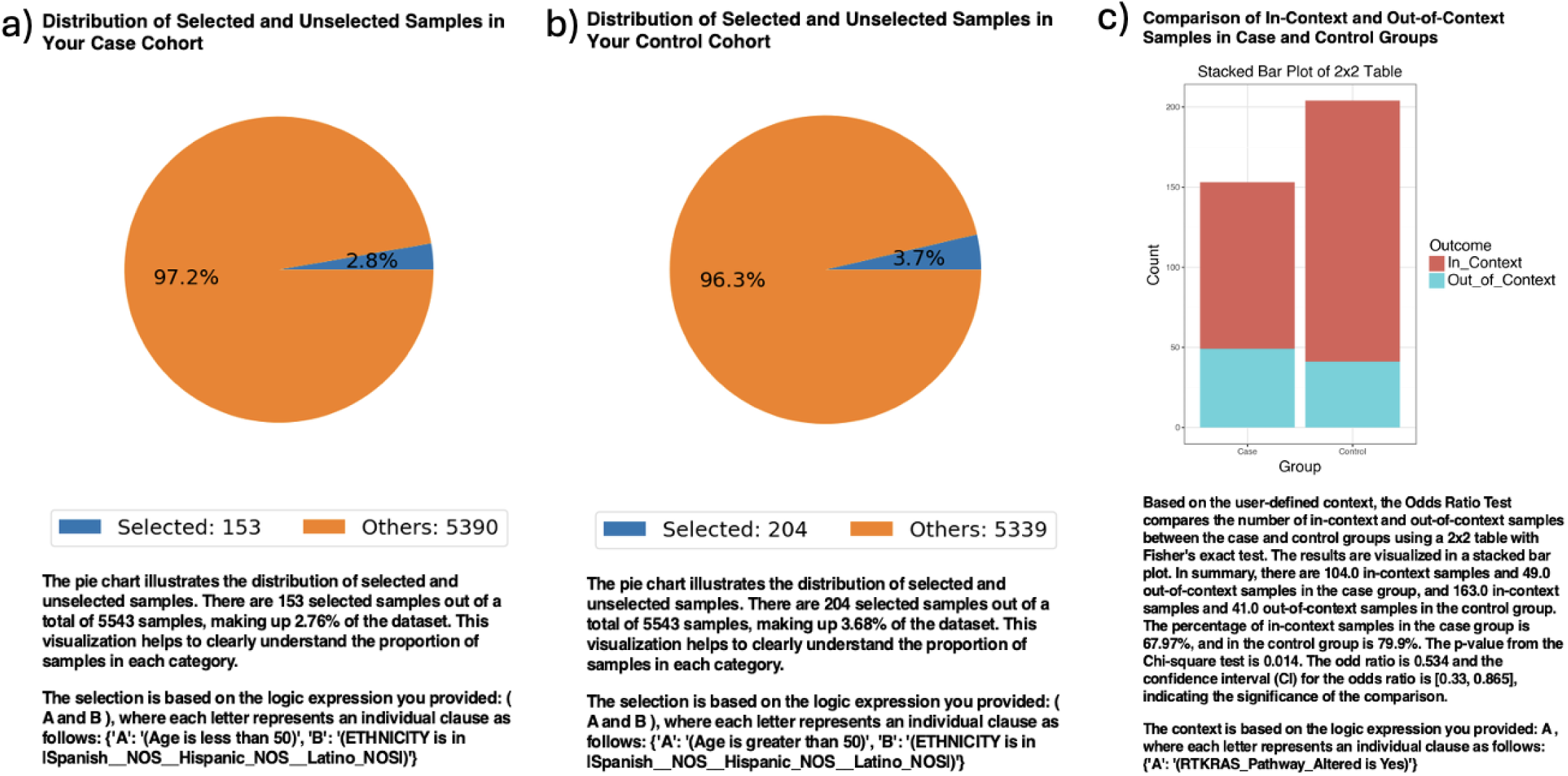
AI-HOPE-RTK-RAS analysis of RTK-RAS pathway alterations in early-vs. late-onset colorectal cancer (CRC) Patients. This figure illustrates the application of AI-HOPE-RTK-RAS to evaluate age-related differences in RTK-RAS pathway alterations among CRC patients using a natural language query and automated odds ratio framework. a) The case cohort includes 153 early-onset CRC (EOCRC) Hispanic/Latino (H/L) patients under the age of 50 (2.8% of the dataset), selected using demographic filters. The pie chart displays the proportion of selected EOCRC H/L patients relative to the full sample population. b) The control cohort consists of 204 late-onset CRC (LOCRC) H/L patients over the age of 50 (3.7% of the dataset), similarly selected by age and ethnicity. A pie chart illustrates their representation within the dataset. c) An odds ratio test compares the frequency of RTK-RAS pathway alterations between the EOCRC and LOCRC H/L cohorts using a 2×2 table and stacked bar plot. RTK-RAS alterations were present in 67.97% of early-onset and 79.9% of late-onset cases. The resulting odds ratio was 0.534 (95% CI: 0.33–0.865, p = 0.014), indicating that early-onset H/L patients were significantly less likely to harbor RTK-RAS alterations than their later-onset counterparts. This result suggests potential differences in molecular pathogenesis by age within this population and underscores AI-HOPE-RTK-RAS’s capacity to support ancestry- and age-stratified genomic analyses through natural language–driven precision oncology.

### Stage-Dependent Outcomes in KRAS-Mutant CRC

Analysis of KRAS-mutant patients receiving Bevacizumab revealed stage-related differences in survival. Patients diagnosed with Stage I–III disease demonstrated significantly better overall survival than those with Stage IV tumors (p = 0.0004), underscoring the prognostic influence of stage and suggesting potential variation in response to targeted therapies based on disease extent (Figure 3).

**Figure 3.**
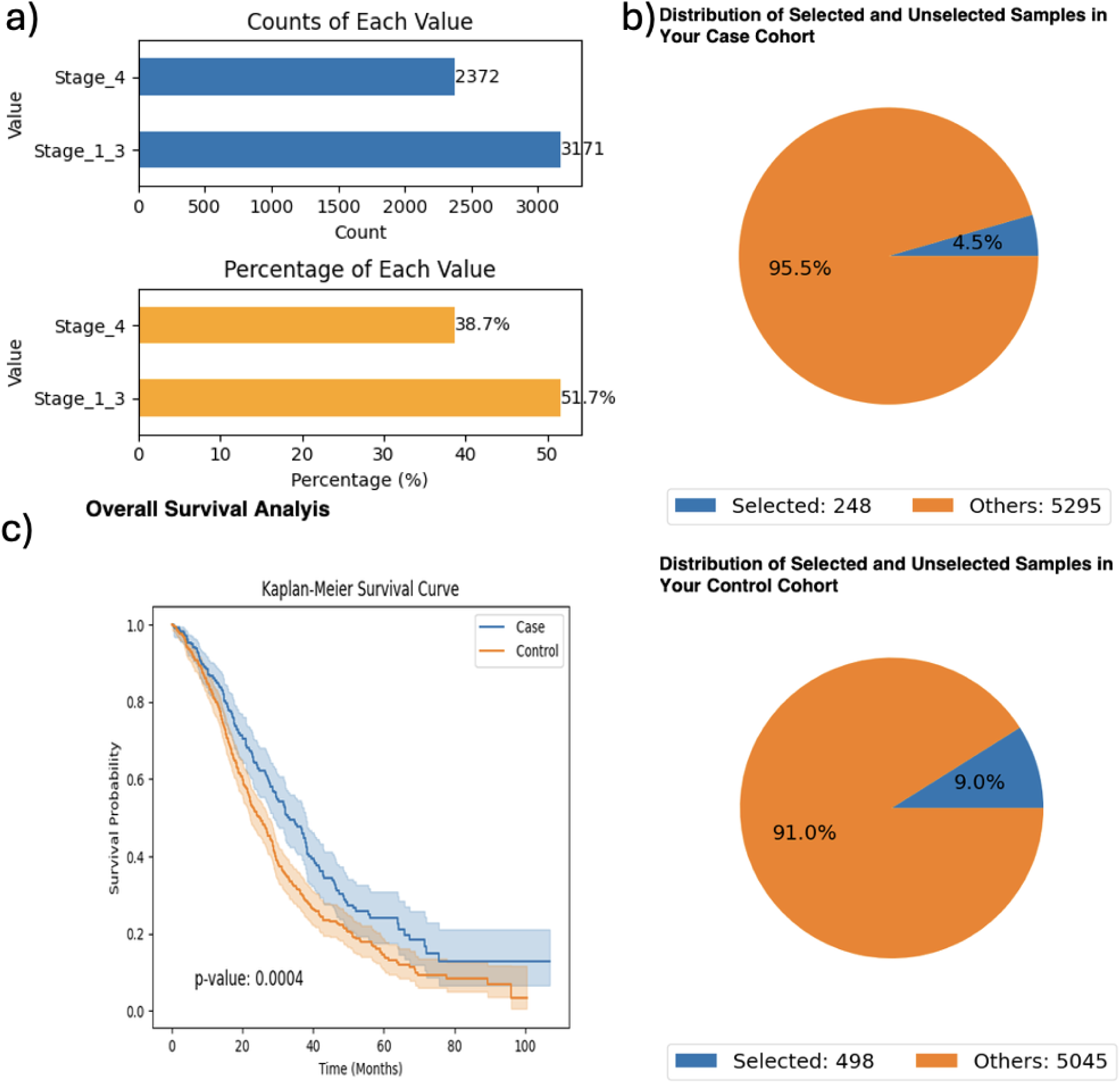
AI-HOPE-RTK-RAS survival analysis of KRAS-mutant colorectal cancer (CRC) patients treated with bevacizumab, stratified by tumor stage. This figure highlights AI-HOPE-RTK-RAS’s capacity to perform integrative survival analysis using clinical, treatment, and mutation-specific variables. The analysis focuses on CRC patients harboring *KRAS* mutations who received Bevacizumab, stratified by tumor stage. a) The exploratory analysis summarizes the distribution of patients across tumor stages. Bar plots show that among *KRAS*-mutant patients treated with Bevacizumab, 3,171 were classified as Stage I–III and 2,372 as Stage IV. Proportionally, Stage I–III represented 51.7% of the cohort, while Stage IV accounted for 38.7%, supporting adequate sample sizes for comparative outcome analysis. b) Pie charts illustrate the subset of patients selected for survival comparison: the case group (Stage I–III) included 248 patients (4.5% of the dataset), while the control group (Stage IV) included 498 patients (9.0%). These visualizations emphasize the relative cohort sizes in the context of the broader population. c) A Kaplan-Meier survival curve compares overall survival between Stage I–III and Stage IV patients within the *KRAS*-mutant, Bevacizumab-treated cohort. The analysis reveals a statistically significant survival advantage for patients with primary-stage disease (Stage I–III), with a p-value of 0.0004. Distinct separation of survival curves and non-overlapping confidence intervals further support the observed outcome disparity. This figure demonstrates AI-HOPE-RTK-RAS’s ability to automate complex stratified survival analyses in a precision oncology context through conversational AI.

### Prognostic Role of MSI in BRAF-Mutant Disease

When evaluating microsatellite status in BRAF-mutant CRC, patients with stable MSI were significantly more likely to have received chemotherapy (OR = 7.226, p < 0.001). Despite this, survival analysis indicated inferior outcomes for the MSI-stable group compared to MSI-instability counterparts (p = 0.0000), highlighting the complexity of MSI-related treatment responses in BRAF-mutant contexts (Figure 4).

**Figure 4.**
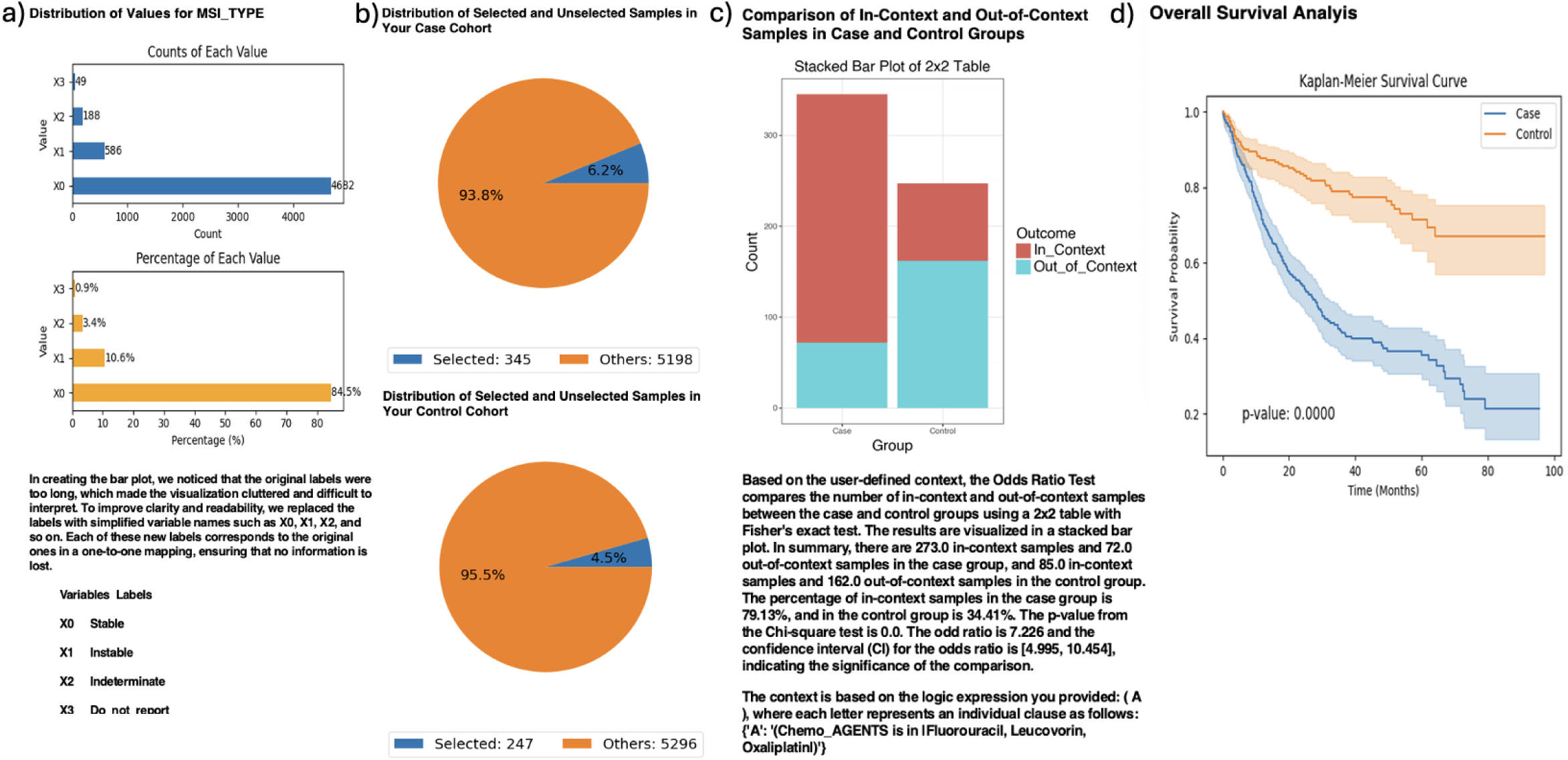
AI-HOPE-RTK-RAS analysis of BRAF-mutant colorectal cancer (CRC) patients by microsatellite stability status. This figure illustrates AI-HOPE-RTK-RAS’s capability to interrogate survival and odds ratio patterns in BRAF-mutant CRC patients stratified by microsatellite stability (MSI) status and chemotherapy exposure. a) The bar charts depict the distribution of MSI types across the dataset. The top panel shows raw counts, with stable MSI (X0) comprising the majority (n = 4,682), followed by instable MSI (X1, n = 586), indeterminate (X2, n = 188), and unknown (X3, n = 49). The bottom chart displays proportions, confirming that stable MSI (84.5%) predominates, with instable MSI accounting for 10.6% of samples. b) Based on the query, pie charts display the relative distribution of selected patients: the case group includes 345 BRAF-mutant CRC patients with stable MSI (6.2% of the dataset), and the control group includes 247 patients with instable MSI (4.5%). These proportions reflect meaningful yet relatively rare molecular subtypes. c) An odds ratio test is performed to evaluate the association between MSI status and chemotherapy exposure (specifically Fluorouracil, Leucovorin, and Oxaliplatin) among BRAF-mutant CRC patients. A stacked bar plot visualizes the outcome (In_Context vs. Out_of_Context). The odds ratio was 7.226 (95% CI: [4.995, 10.454], *p* < 0.001), indicating a significantly higher likelihood of chemotherapy use among patients with stable MSI compared to those with instable MSI. d) Kaplan-Meier survival analysis compares overall survival between the two groups. The curve shows significantly worse outcomes for the stable MSI cohort relative to the instable MSI cohort (*p* = 0.0000), with early divergence of survival probabilities and non-overlapping confidence intervals. This result underscores the prognostic relevance of MSI status in BRAF-mutant CRC and highlights AI-HOPE-RTK-RAS’s power in generating clinical-genomic hypotheses using real-world datasets.

### Impact of RTK-RAS Alterations on EOCRC Treated with FOLFOX

In patients with EOCRC treated with FOLFOX, the presence of RTK-RAS pathway alterations was associated with worse survival (p = 0.0262). However, ethnic distribution between groups was statistically similar (OR = 1.00, p = 1.00), supporting the general applicability of this finding across the EOCRC H/L population (Figure 5).

**Figure 5.**
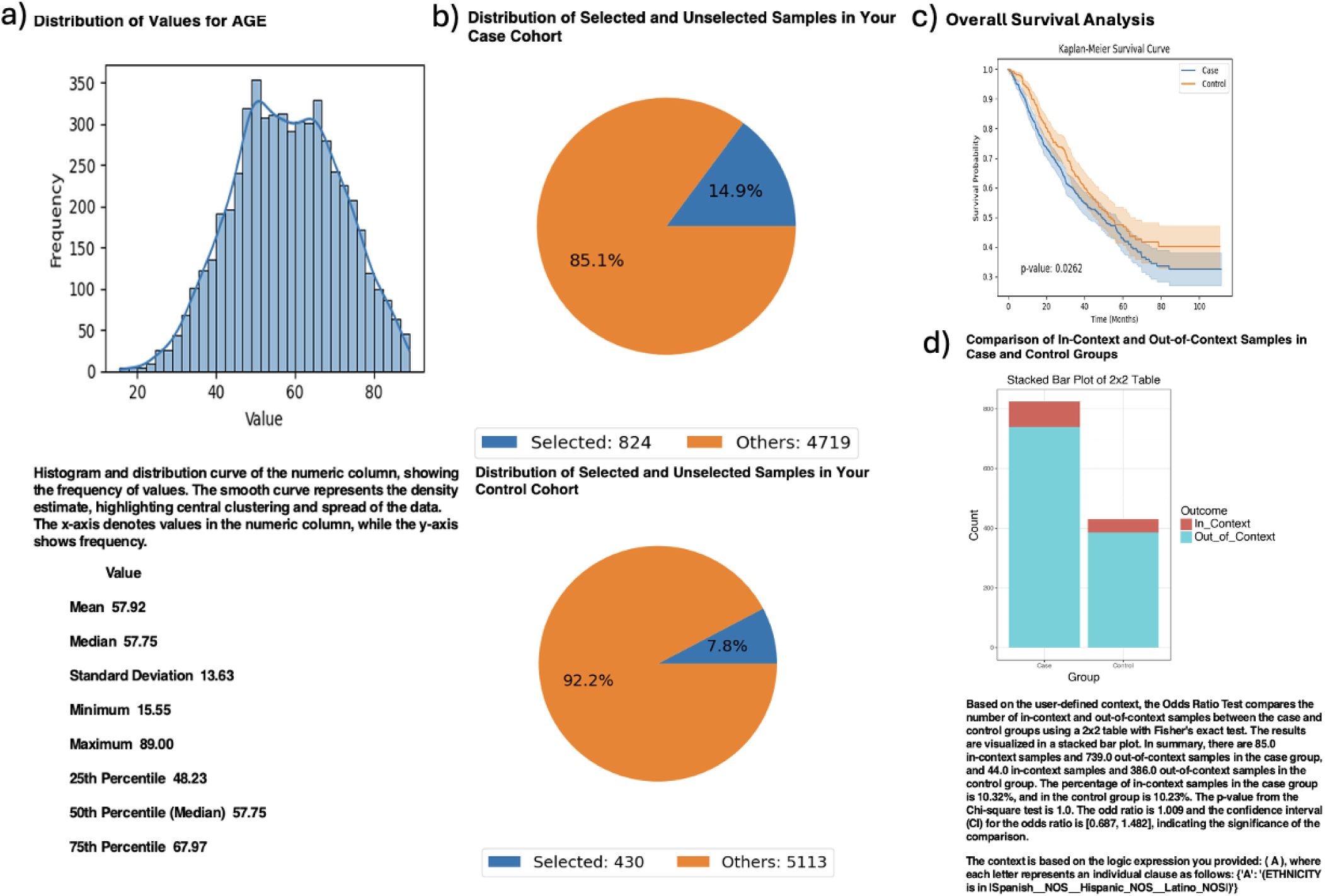
AI-HOPE-RTK-RAS analysis of early-onset colorectal cancer (CRC) patients treated with FOLFOX stratified by RTK-RAS pathway alteration. This figure presents the results of a natural language query performed using AI-HOPE-RTK-RAS, examining the clinical and genomic impact of RTK-RAS pathway alterations in early-onset colorectal cancer (EOCRC) patients of Hispanic/Latino (H/L) ancestry treated with FOLFOX (Fluorouracil, Leucovorin, Oxaliplatin). a) A histogram displays the distribution of patient ages across the dataset, with a smooth density curve highlighting the central tendency and spread. The mean age is 57.92 years, and the median is 57.75 years, confirming the appropriateness of using <50 years as a cutoff to define EOCRC. b) Pie charts show the relative sample sizes of the defined cohorts. The case group includes 824 patients under age 50 with RTK-RAS pathway alterations and FOLFOX treatment (14.9% of the dataset), while the control group consists of 430 EOCRC patients without RTK-RAS alterations who also received FOLFOX (7.8%). These visualizations reflect the selective filtering enabled through AI-HOPE’s natural language-driven interface. c) Kaplan-Meier survival curves compare overall survival between the case and control groups. Although both cohorts were treated with FOLFOX, patients with RTK-RAS pathway alterations showed moderately worse survival, with a p-value of 0.0262. The divergence of the curves and non-overlapping confidence intervals suggest a potential prognostic effect of RTK-RAS alterations in early-onset Hispanic/Latino CRC. d) An odds ratio test was conducted to examine the enrichment of Hispanic/Latino ethnicity among cases versus controls. The stacked bar plot illustrates a 2×2 comparison of in-context (H/L) and out-of-context (non-Hispanic Whites-NHW) patients. The odds ratio was 1.00 (95% CI: [0.687, 1.482], p = 1.00), indicating no significant enrichment of Hispanic/Latino individuals in either group. This analysis underscores AI-HOPE-RTK-RAS’s ability to execute multifaceted queries integrating clinical, genomic, and demographic variables to support precision oncology research.

### Ancestry-Specific Mutation Enrichment

The platform revealed enrichment of several noncanonical RTK-RAS alterations in early-onset disease. CBL mutations were nearly five times more common in EOCRC H/L versus LOCRC H/L patients (OR = 4.842, p = 0.071; Figure S1), and NF1 mutations showed statistically significant overrepresentation (OR = 2.53, p = 0.045; Figure S2). Comparative analyses between H/L and Non-Hispanic White EOCRC patients confirmed higher frequencies of MAPK3 (OR = 4.26, p = 0.043; Figure S3), CBL (OR = 4.07, p = 0.005; Figure S4), and NF1 mutations (OR = 2.06, p = 0.021; Figure S5) in the Hispanic/Latino subgroup.

### Tumor Location, Sex, and Prognosis in KRAS- and NF1-Mutated CRC

Further interrogation of KRAS-mutated CRC revealed no significant difference in female representation (OR = 0.985, p = 0.955) or survival (p = 0.7774) between proximal and distal tumor locations (Figure S6). In contrast, patients with NF1-mutated primary CRC tumors experienced significantly improved survival compared to NF1 wild-type counterparts (p = 0.0000), suggesting a possible protective or distinct molecular role of NF1 in early-stage CRC biology (Figure S7).

### Additional Findings and Analytical Capabilities

Beyond hypothesis-driven queries, AI-HOPE-RTK-RAS facilitated exploratory investigation of mutation co-occurrence, therapeutic context, and demographic interaction effects. For example, the platform enabled comparisons of chemotherapy exposure across MSI subtypes, as well as associations between sex, stage, and survival in genetically defined subgroups. The system’s ability to map biological hypotheses onto real-world datasets through conversational prompts accelerated pattern discovery and hypothesis generation.

AI-HOPE-RTK-RAS also demonstrated strengths in interpretability and transparency, providing structured visual summaries (e.g., pie charts, bar plots, survival curves) alongside contextual narrative descriptions. These outputs reduced the analytical barrier for researchers without programming expertise and allowed for quick iteration of refined queries.

## Discussion

The development of AI-HOPE-RTK-RAS marks a significant advancement in precision oncology, offering a conversational artificial intelligence platform tailored to interrogate RTK-RAS pathway alterations in CRC. By leveraging large language models to enable natural language interaction with genomic and clinical datasets, the system addresses critical limitations of traditional bioinformatics tools, including static interfaces and restricted accessibility for non-specialists.

Our findings underscore AI-HOPE-RTK-RAS’s capacity to replicate known associations while uncovering novel molecular and clinical patterns, particularly among EOCRC and high-risk populations. The reduced prevalence of canonical RTK-RAS mutations in EOCRC H/L patients compared to later-onset cohorts (OR = 0.534, p = 0.014) aligns with recent reports suggesting distinct etiologic and molecular profiles in younger patients. These differences highlight the importance of age- and ancestry-stratified analyses to avoid misclassification of risk and misapplication of therapeutic strategies.

Survival analyses further demonstrated AI-HOPE-RTK-RAS’s capability to delineate prognostic variation across genetically defined subgroups. For instance, among KRAS-mutant patients treated with Bevacizumab, those with Stage I–III disease had significantly better outcomes than those with Stage IV tumors (p = 0.0004), emphasizing the prognostic impact of stage even within mutation-defined cohorts. Additionally, our findings in BRAF-mutant CRC showed that MSI-stable patients were more likely to receive chemotherapy (OR = 7.226) but experienced worse survival than MSI-instable counterparts (p = 0.00001), consistent with prior literature linking MSI status to immunogenicity and treatment response.

Importantly, AI-HOPE-RTK-RAS revealed several ancestry-specific mutation patterns that warrant further exploration. Enrichment of noncanonical RTK-RAS genes— including CBL (OR = 4.07, p = 0.005), MAPK3 (OR = 4.26, p = 0.043), and NF1 (OR = 2.06, p = 0.021)—in H/L EOCRC patients underscores the need to broaden biomarker discovery efforts beyond canonical alterations such as KRAS and BRAF [5]. These findings are consistent with recent efforts to characterize high-risk populations in cancer genomics, which have revealed divergent molecular signatures with implications for targeted therapy and biomarker development.

AI-HOPE-RTK-RAS also supported exploration of sex, tumor location, and survival outcomes in KRAS- and NF1-mutant contexts. While proximal and distal KRAS-mutant tumors did not differ significantly in survival or sex representation (p = 0.7774; p = 0.955), NF1-mutated primary CRC cases demonstrated significantly better survival than NF1 wild-type tumors (p = 0.00001), suggesting NF1 may serve as a favorable prognostic marker in early-stage disease. This is aligned with prior work implicating NF1 in modulating MAPK signaling and tumor suppressor functions in various cancers [5,41–43].

Beyond hypothesis testing, AI-HOPE-RTK-RAS enabled exploratory data analysis through iterative, language-driven workflows that required no coding. This functionality proved particularly valuable in generating real-time insights into co-mutation frequency, chemotherapy stratification, and context-aware survival differences. By unifying genomic and clinical data into a conversational interface, the platform lowered barriers for precision oncology research and empowered users to pursue complex analyses previously requiring substantial computational expertise.

Nevertheless, certain limitations merit discussion. First, our analyses were restricted to publicly available datasets, which may underrepresent minority populations or lack detailed clinical annotations. Second, while the natural language interface is a major strength, it relies on accurate query interpretation and context parsing, which may introduce challenges for ambiguous or nested questions. Continued validation against curated benchmarks and expanded integration with electronic health records will be essential to improving generalizability and clinical adoption.

## Conclusion

In conclusion, AI-HOPE-RTK-RAS provides a robust, scalable platform to investigate RTK-RAS biology in CRC, with applications in biomarker discovery, cancer genetics, and treatment response stratification. By coupling AI-driven querying with real-time, multimodal data integration, the system offers a new paradigm for accessible, interactive precision oncology research. Future work will focus on extending AI-HOPE capabilities to additional pathways, longitudinal datasets, and clinical decision support environments.

## Data Availability

All data used in the present study is publicly available at https://www.cbioportal.org/ and https://genie.cbioportal.org. Additional data can be provided upon reasonable request to the authors.

---

**Figure S1.**
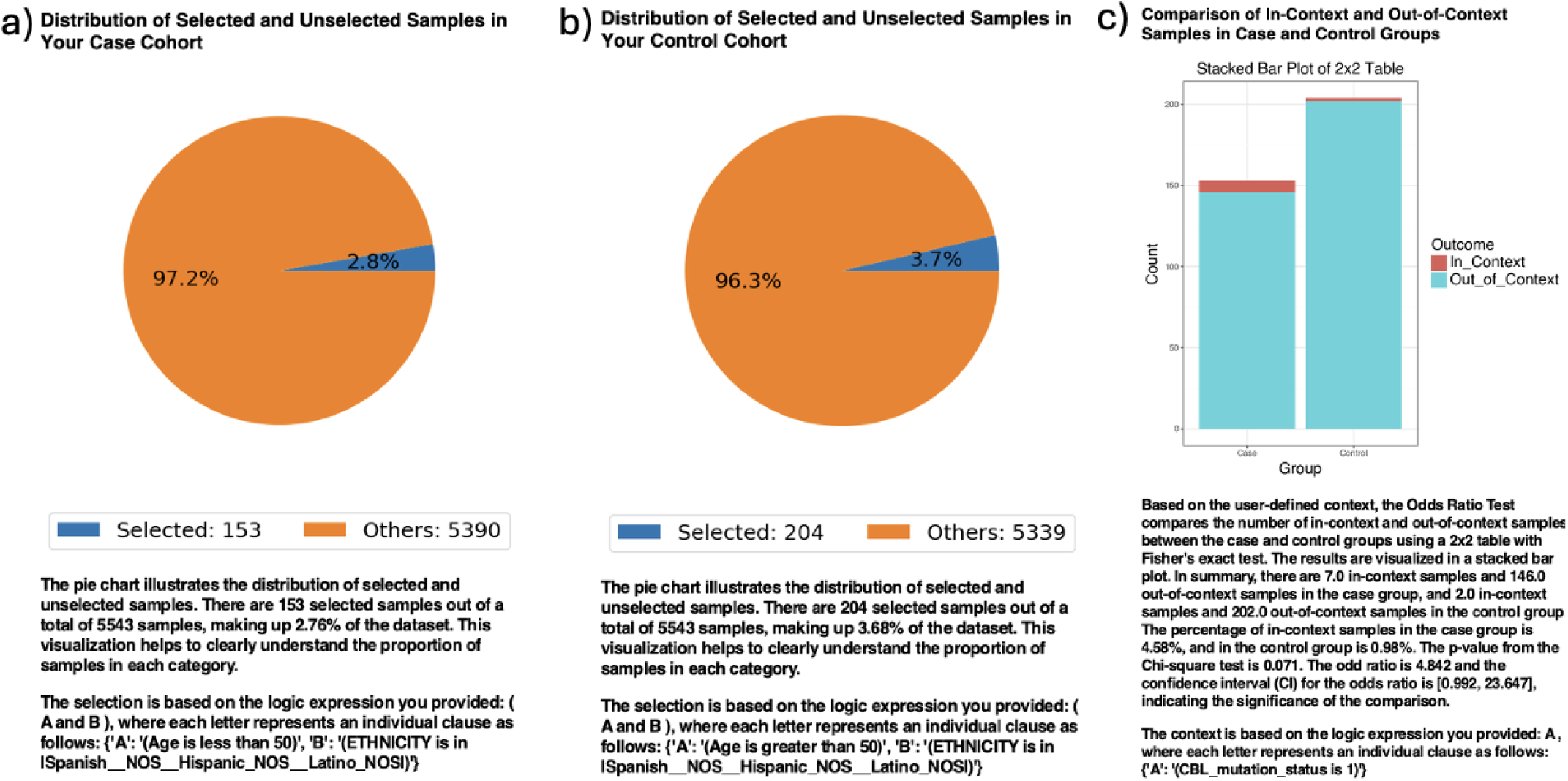
AI-HOPE-RTK-RAS analysis of CBL mutation frequency in early-onset versus late-onset colorectal cancer (CRC) patients. This figure illustrates AI-HOPE-RTK-RAS’s capacity to evaluate gene-specific alterations, in this case focusing on CBL mutations, within demographically defined CRC cohorts. The platform was used to compare the prevalence of CBL mutations in early-onset CRC (EOCRC) versus late-onset CRC (LOCRC) Hispanic/Latino (H/L) patients using a natural language–driven odds ratio framework. a) The case cohort includes 153 EOCRC H/L patients under the age of 50 (2.8% of the dataset), identified using age and ethnicity filters. A pie chart visualizes the proportion of selected EOCRC cases relative to the full dataset. b) The control cohort consists of 204 H/L patients over the age of 50 with LOCRC (3.7% of the dataset). The pie chart shows their representation within the population. c) An odds ratio test compares the presence of CBL mutations across both groups using a 2×2 contingency table and stacked bar plot. CBL mutations were observed in 4.58% of EOCRC and 0.98% of LOCRC samples. The resulting odds ratio was 4.842 (95% CI: 0.992–23.647, p = 0.071), indicating a nearly fivefold increase in CBL mutation odds in early-onset cases, though the result did not reach conventional statistical significance. This trend highlights a potential enrichment of CBL mutations in EOCRC H/L patients and demonstrates AI-HOPE-RTK-RAS’s utility for uncovering subgroup-specific genomic patterns that may warrant further investigation in larger cohorts.

**Figure S2.**
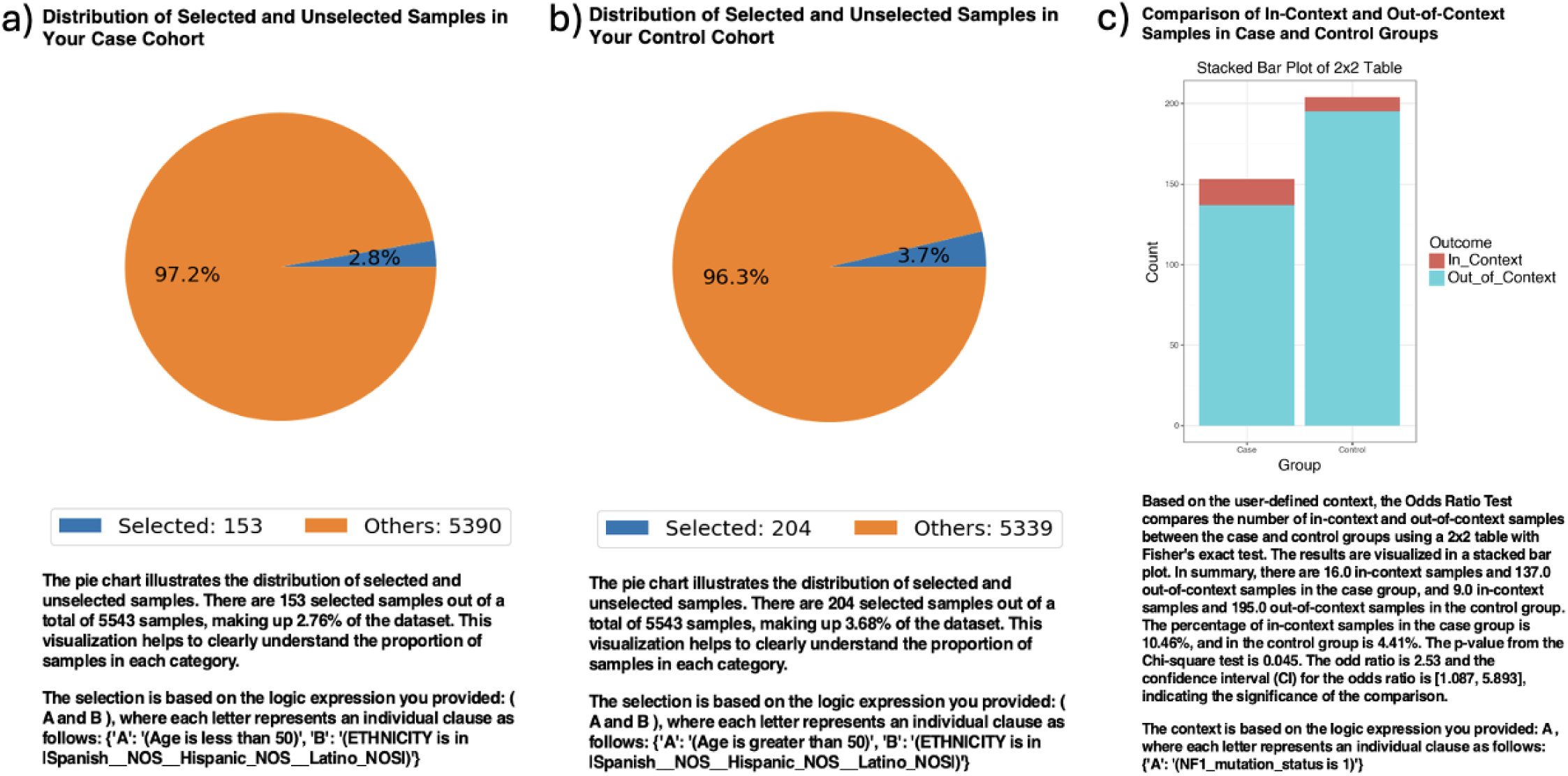
AI-HOPE-RTK-RAS evaluation of NF1 mutation frequency in early-onset versus late-onset colorectal cancer (CRC) Patients. This figure demonstrates AI-HOPE-RTK-RAS’s functionality in detecting age-related genomic differences through natural language–driven analysis, focusing on *NF1* mutation prevalence among Hispanic/Latinos (H/L) CRC patients. a) The case cohort includes 153 EOCRC H/L patients under the age of 50 (2.8% of the total dataset), identified using filters for both age and ethnicity. A pie chart illustrates the proportion of selected EOCRC H/L patients relative to the overall CRC cohort. b) The control cohort consists of 204 LOCRC H/L patients over the age of 50 (3.7% of the dataset), selected using the same ethnicity criteria. A pie chart shows this group’s relative representation. c) An odds ratio test was used to compare the frequency of *NF1* mutations between the EOCRC and LOCRC groups. The bar plot displays a 2×2 comparison of *NF1*-mutated (in-context) versus wild-type (out-of-context) samples. *NF1*mutations were observed in 10.46% of EOCRC samples and 4.41% of LOCRC samples. The odds ratio was 2.53 (95% CI: [1.087, 5.893], *p* = 0.045), indicating that early-onset patients had more than twice the odds of harboring *NF1* mutations compared to their later-onset counterparts. This statistically significant difference supports a potential role for *NF1*alterations in the molecular landscape of early-onset CRC among H/L populations and highlights the utility of AI-HOPE-RTK-RAS for uncovering clinically relevant, subgroup-specific genomic patterns.

**Figure S3.**
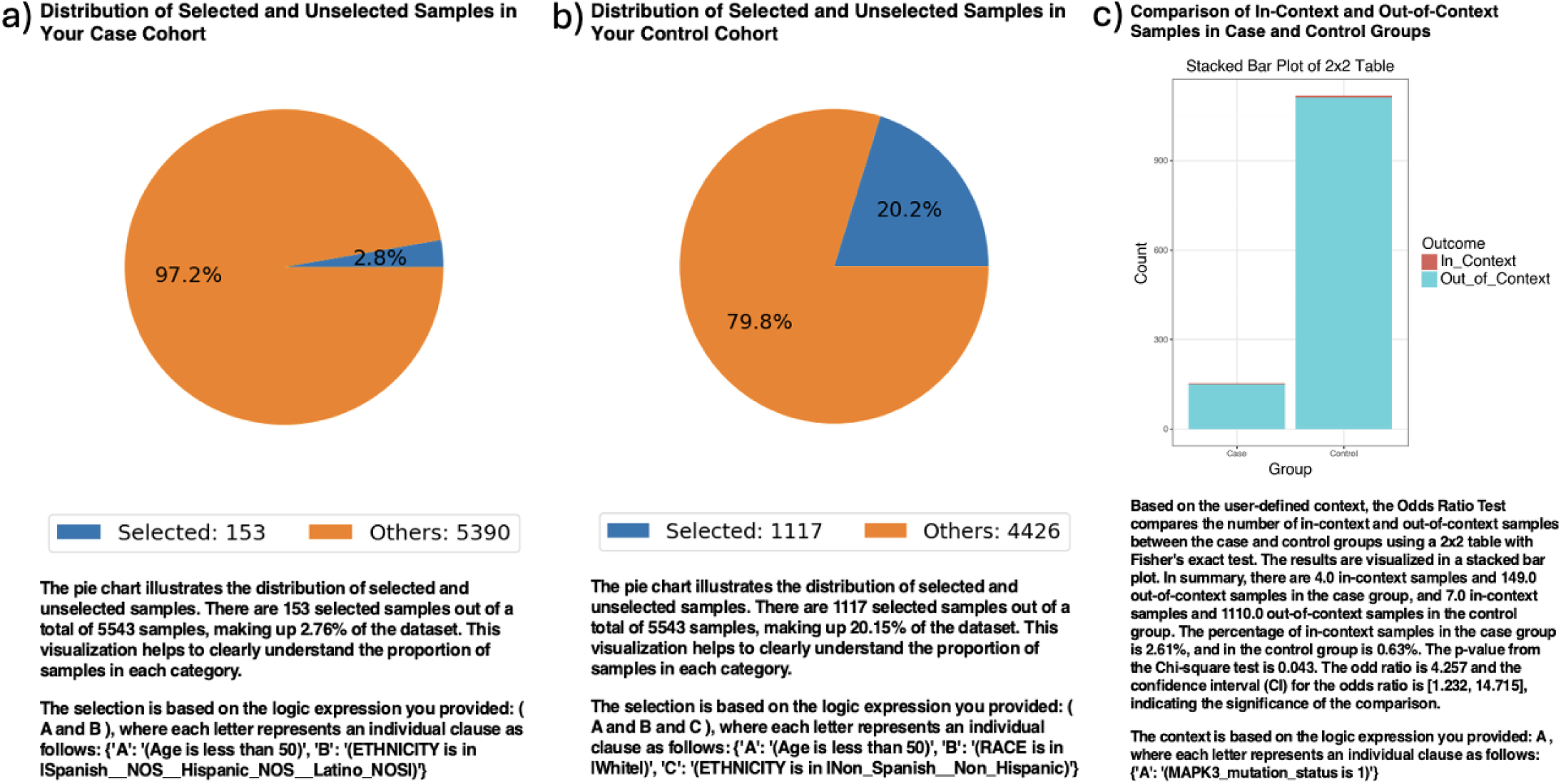
AI-HOPE-RTK-RAS analysis of MAPK3 mutation frequency in early-onset colorectal cancer (CRC) patients by ethnicity. This figure illustrates AI-HOPE-RTK-RAS’s ability to identify ethnicity-associated genomic alterations through natural language–driven querying, specifically focusing on *MAPK3* mutations in early-onset colorectal cancer (EOCRC) patients. a) The case cohort includes 153 EOCRC Hispanic/Latino (H/L) patients under the age of 50 (2.8% of the dataset), selected using demographic filters based on ethnicity. A pie chart displays the proportion of EOCRC HL patients among all samples. b) The control cohort consists of 1,117 EOCRC Non-Hispanic White (NHW) patients under age 50 (20.2% of the dataset), identified based on combined race and ethnicity filters. A corresponding pie chart visualizes this group’s relative dataset representation. c) An odds ratio test compares the frequency of *MAPK3* mutations between the two cohorts. As shown in the stacked bar plot, *MAPK3* mutations were detected in 2.61% of EOCRC HL samples and 0.63% of EOCRC NHW samples. The odds ratio was 4.26 (95% CI: [1.232, 14.715], *p* = 0.043), indicating that H/L EOCRC patients had more than four times the odds of harboring a *MAPK3* mutation compared to their NHW counterparts. This statistically significant finding highlights a potential enrichment of *MAPK3* mutations in the Hispanic/Latino early-onset CRC population and supports the need for further investigation into *MAPK3* as a possible biomarker or driver of ethnicity-specific molecular differences in EOCRC.

**Figure S4.**
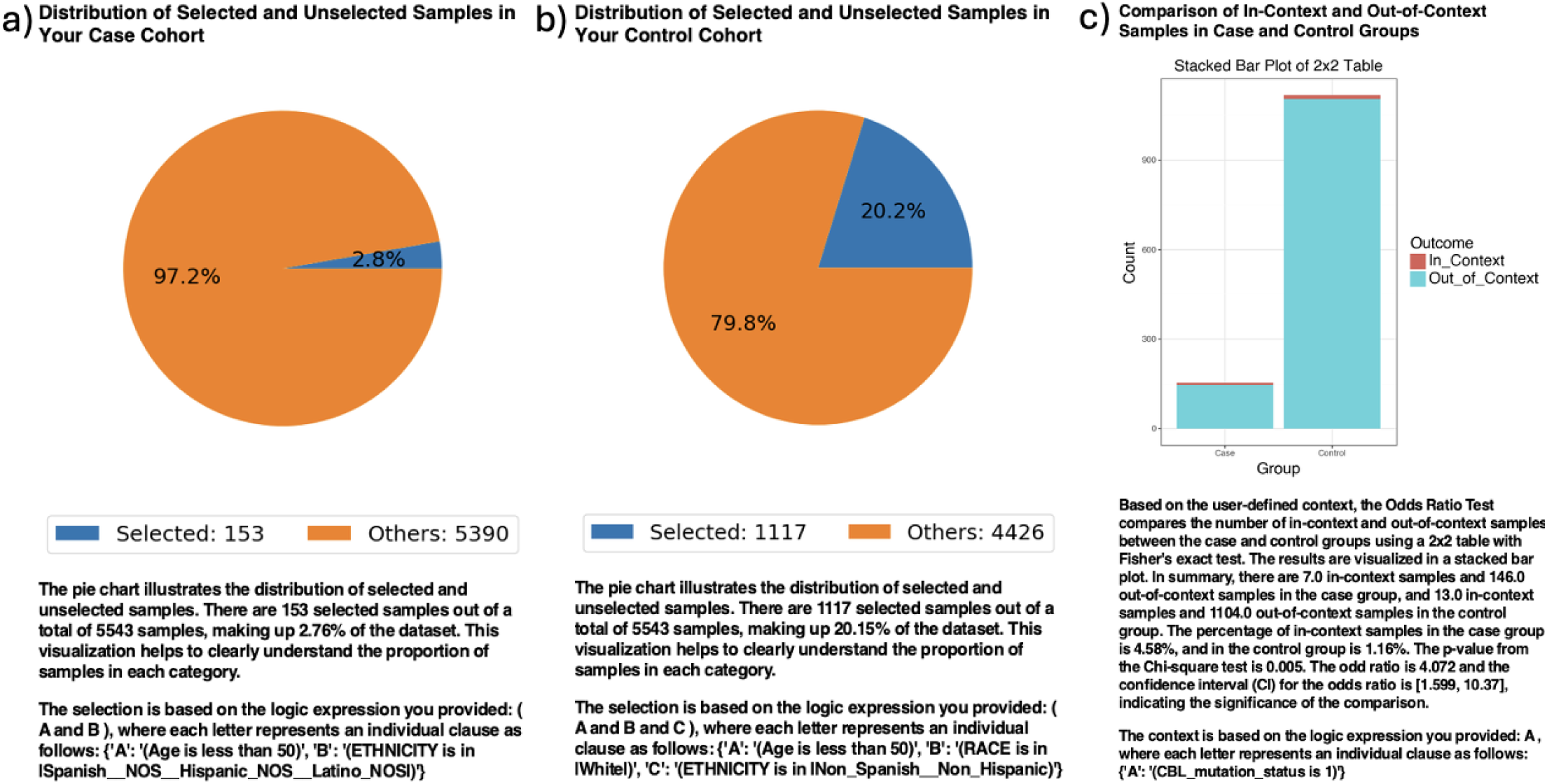
AI-HOPE-RTK-RAS analysis of CBL mutation frequency in early-onset colorectal cancer (EOCRC) patients by ethnicity. This figure demonstrates AI-HOPE-RTK-RAS’s capability to identify ancestry-related mutation patterns through natural language–driven analysis, focusing on CBL mutations in EOCRC patients. a) The case cohort consists of 153 Hispanic/Latino (H/L) EOCRC patients under the age of 50 (2.8% of the dataset), selected using ethnicity-specific filters. A pie chart illustrates the proportion of selected cases within the total CRC dataset. b) The control cohort includes 1,117 Non-Hispanic White (NHW) EOCRC patients under age 50 (20.2% of the dataset), identified through combined race and ethnicity filters. A corresponding pie chart displays their relative representation. c) An odds ratio test compares the frequency of CBL mutations between the two groups. As visualized in the stacked bar plot, CBL mutations were present in 4.58% of EOCRC H/L cases and 1.16% of EOCRC NHW cases. The calculated odds ratio was 4.07 (95% CI: [1.599, 10.37], p = 0.005), indicating that H/L EOCRC patients had more than four times the odds of harboring a CBL mutation compared to NHW counterparts. This statistically significant enrichment suggests that CBL may play a more prominent role in the molecular landscape of early-onset CRC in H/L patients and supports its potential relevance as a biomarker or oncogenic driver in this population.

**Figure S5.**
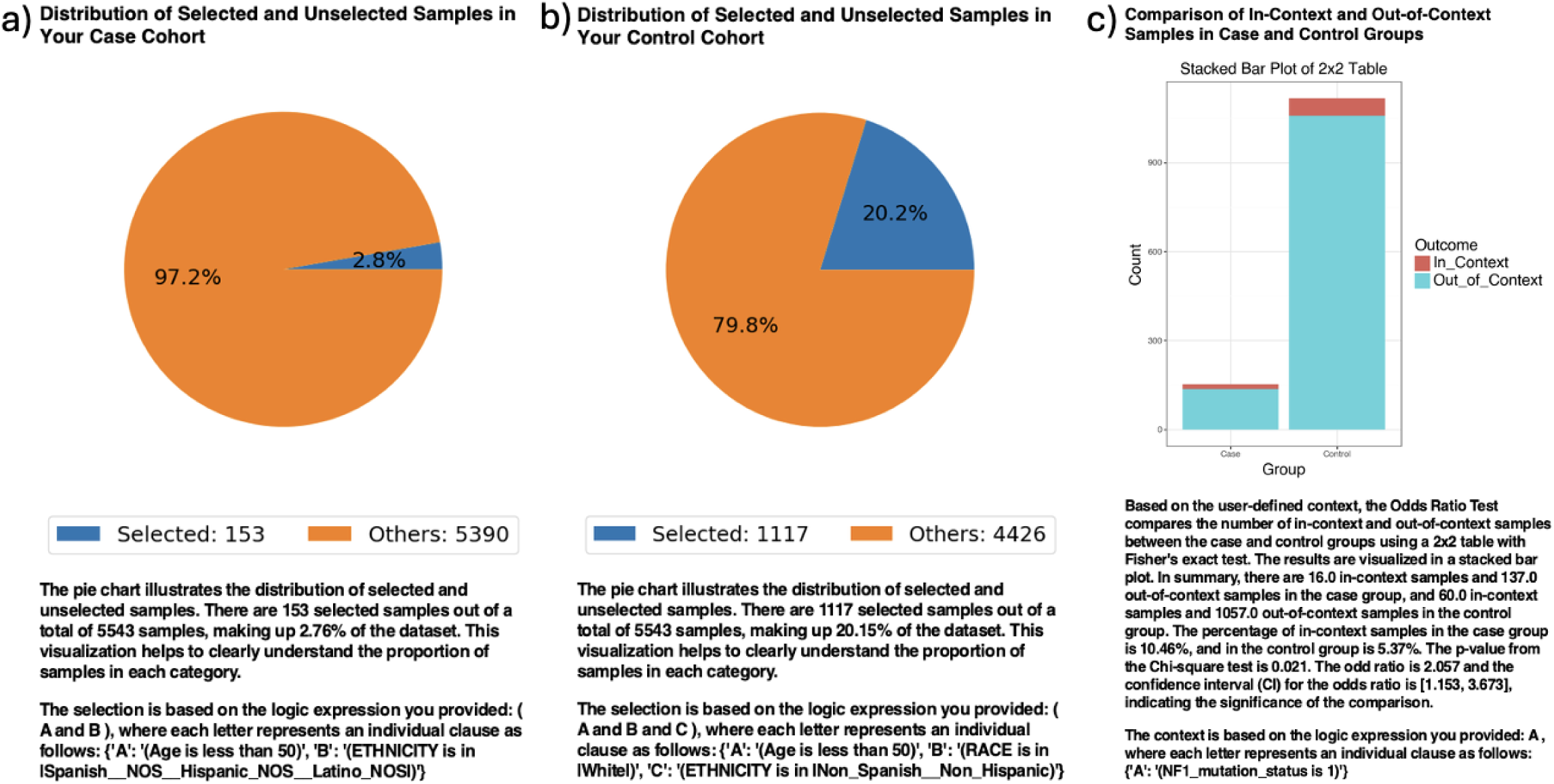
AI-HOPE-RTK-RAS analysis of NF1 mutation frequency in early-onset colorectal cancer (EOCRC) patients by ethnicity. This figure demonstrates AI-HOPE-RTK-RAS’s capability to uncover ancestry-associated genomic alterations using natural language–driven queries, focusing on *NF1* mutation prevalence in early-onset colorectal cancer (EOCRC). a) The case cohort includes 153 Hispanic/Latino (H/L) EOCRC patients under the age of 50 (2.8% of the dataset), selected based on ethnicity filters. A pie chart illustrates the proportion of selected H/L patients within the overall cohort. b) The control cohort consists of 1,117 Non-Hispanic White (NHW) EOCRC patients under the age of 50 (20.2% of the dataset), identified through race and ethnicity filtering. The pie chart reflects the representation of this group in the dataset. c) An odds ratio test evaluates the frequency of *NF1* mutations between the two cohorts. The stacked bar plot visualizes the comparison of in-context (*NF1*-mutated) versus out-of-context samples in both groups. *NF1* mutations were found in 10.46% of EOCRC HL samples and 5.37% of EOCRC NHW samples. The odds ratio was 2.06 (95% CI: [1.153, 3.673], *p*= 0.021), indicating that Hispanic/Latino patients had approximately double the odds of harboring an *NF1* mutation compared to NHW patients. This statistically significant enrichment suggests that *NF1* may play a more prominent role in the tumor biology of early-onset CRC in Hispanic/Latino populations and highlights AI-HOPE-RTK-RAS’s utility in conducting ancestry-aware, gene-specific analyses to inform precision oncology.

**Figure S6.**
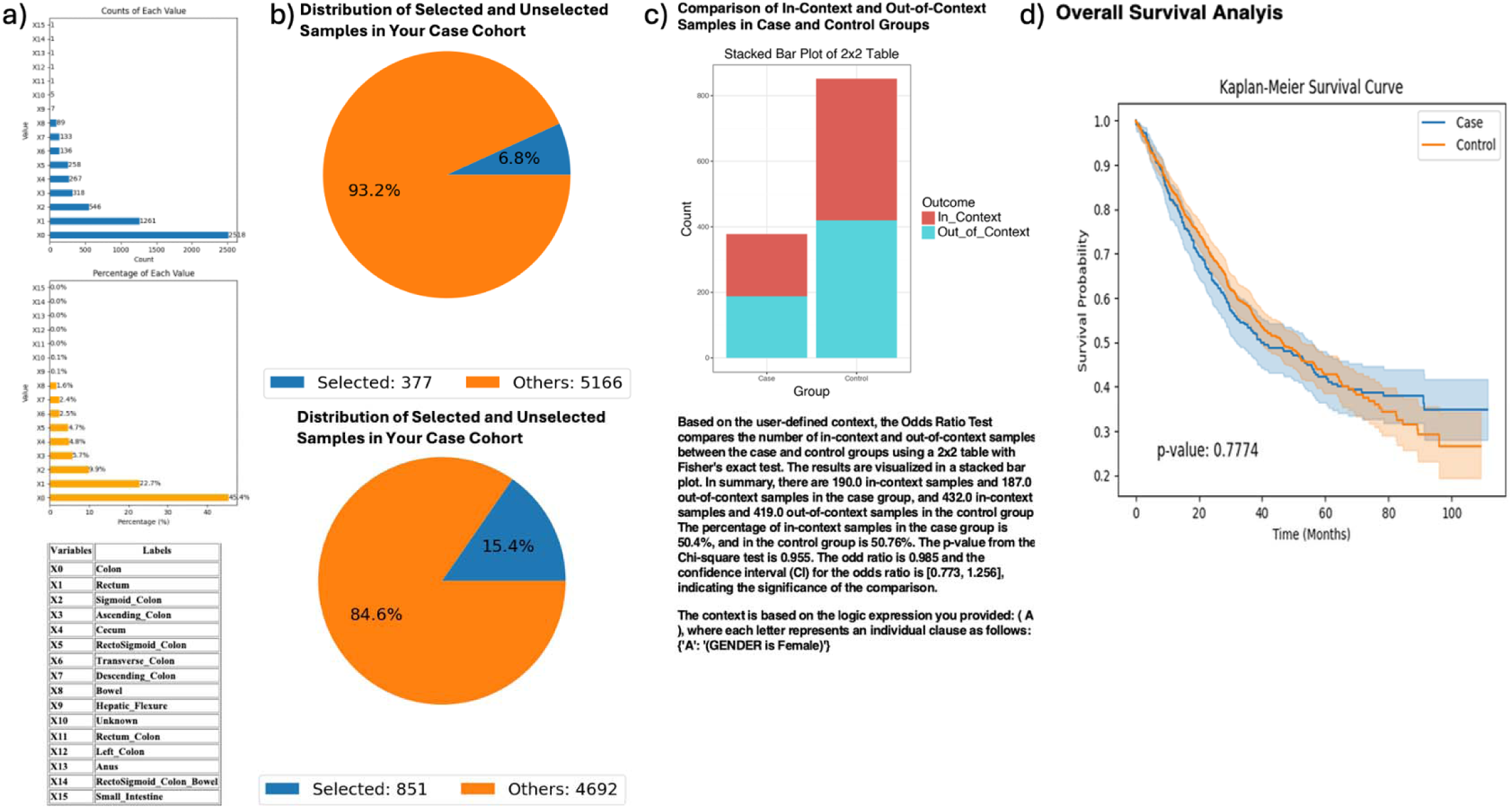
AI-HOPE-RTK-RAS analysis of colorectal cancer (CRC) tumors with KRAS mutations by tumor location: proximal vs. distal colon. This figure demonstrates the use of AI-HOPE-RTK-RAS to compare clinical outcomes and demographic patterns in KRAS-mutated CRC patients based on tumor location— proximal (case group) versus distal (control group)—with additional odds ratio analysis for sex-based enrichment. a) The case and control groups are defined by primary tumor site. The bar charts show the frequency and proportion of CRC primary sites across the dataset. Proximal sites (e.g., Cecum, Ascending Colon, Transverse Colon) are grouped as the case cohort, while distal sites (e.g., Sigmoid Colon, Rectum) form the control. Colon (X0), Sigmoid Colon (X2), and Ascending Colon (X3) are among the most frequently represented locations, with the majority of tumors arising in the distal colon. b) Pie charts illustrate the number of selected samples from each cohort relative to the dataset. The proximal (case) cohort includes 377 KRAS-mutant samples (6.8%), while the distal (control) cohort includes 851 samples (15.4%). This visualization highlights the relatively lower frequency of proximal tumors with KRAS mutations in the analyzed dataset. c) An odds ratio test evaluates sex-based enrichment by comparing the proportion of female patients between cohorts. The stacked bar chart shows the number of in-context (female) and out-of-context (non-female) samples for both groups. The odds ratio was 0.985 (95% CI: [0.773, 1.256], *p* = 0.955), indicating no significant difference in female representation between proximal and distal KRAS-mutant tumor sites. d) Kaplan-Meier survival curves compare overall survival between proximal and distal CRC patients with KRAS mutations. While the curves appear similar and intersect at later timepoints, the survival difference is not statistically significant (*p* = 0.7774), suggesting comparable outcomes across tumor locations in this KRAS-mutant subgroup. Confidence intervals are shown for both groups, confirming the robustness of this observation.

**Figure S7.**
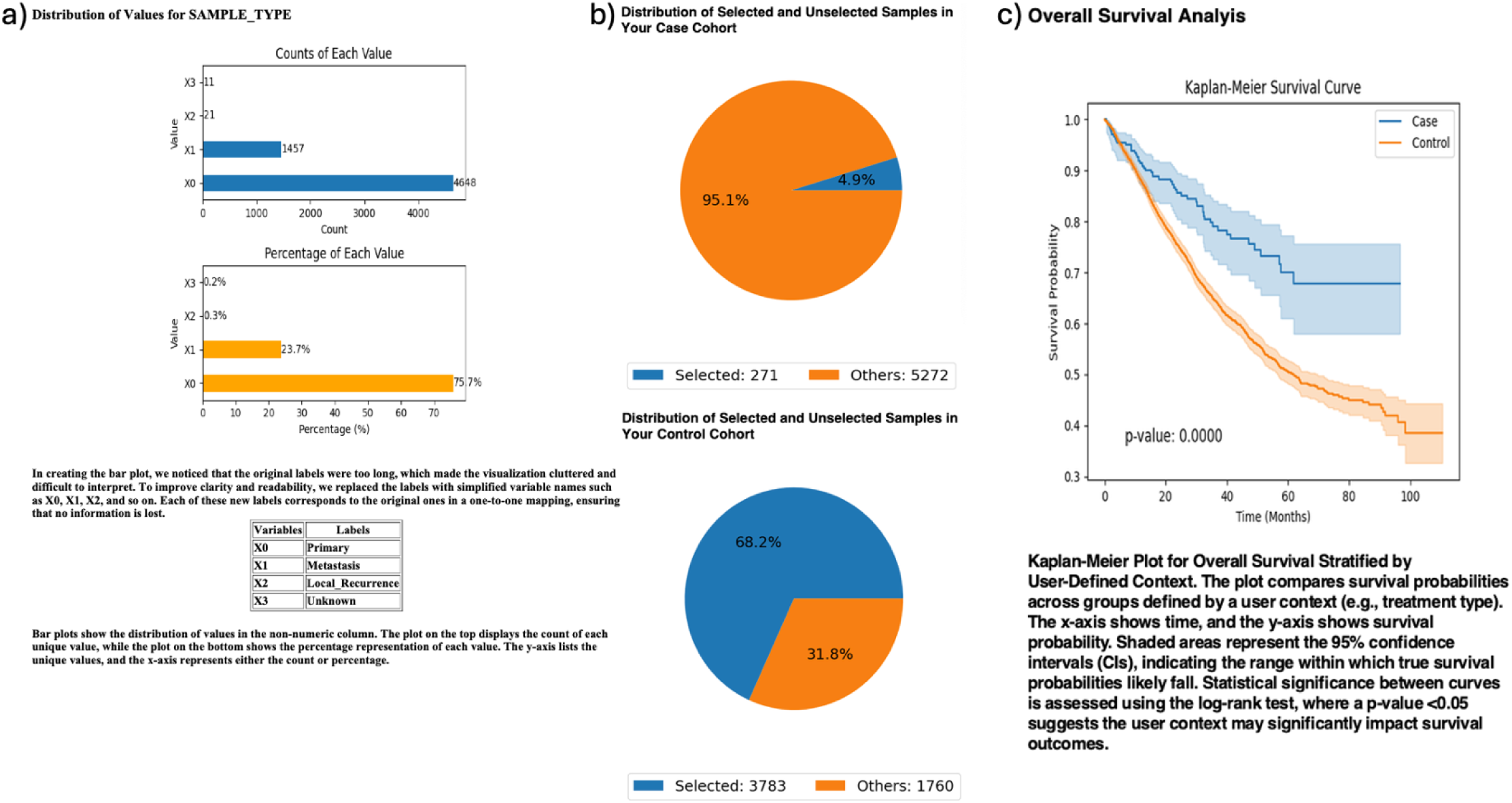
AI-HOPE-RTK-RAS analysis of primary colorectal cancer (CRC) tumors with and without NF1 mutations. This figure showcases AI-HOPE-RTK-RAS’s ability to explore clinical outcomes in colorectal cancer (CRC) patients by comparing primary tumors with and without *NF1* mutations. a) The analysis begins by filtering for primary tumor samples within the CRC dataset. The bar charts summarize the distribution of tumor sample types. Primary tumors (X0) represent the majority (n = 4,648; 75.7%), followed by metastases (X1; 23.7%) and rare classifications such as local recurrence and unknown. This selection ensures that the subsequent comparison focuses solely on primary CRC. b) Two cohorts are generated based on *NF1* mutation status. The case cohort includes 271 primary CRC tumors with *NF1*mutations (4.9%), while the control cohort includes 3,783 primary tumors without *NF1* mutations (68.2%). Pie charts visually represent the selected subsets relative to the entire dataset, illustrating the difference in mutation frequency across the primary tumor landscape. c) Kaplan-Meier survival curves compare overall survival between the *NF1*-mutated and wild-type groups. The survival difference is statistically significant (p = 1 × 10⁻□), with the *NF1*-mutated cohort showing a markedly improved survival probability over time. Confidence intervals are shaded to reflect statistical robustness. These results suggest that *NF1* mutations may confer favorable prognostic impact in primary CRC and highlight the utility of AI-HOPE-RTK-RAS in facilitating rapid, mutation-specific survival analyses across clinically relevant subgroups.

